# Sequencing identifies multiple early introductions of SARS-CoV-2 to the New York City Region

**DOI:** 10.1101/2020.04.15.20064931

**Authors:** Matthew T. Maurano, Sitharam Ramaswami, Paul Zappile, Dacia Dimartino, Ludovic Boytard, André M. Ribeiro-dos-Santos, Nicholas A. Vulpescu, Gael Westby, Guomiao Shen, Xiaojun Feng, Megan S. Hogan, Manon Ragonnet-Cronin, Lily Geidelberg, Christian Marier, Peter Meyn, Yutong Zhang, John Cadley, Raquel Ordoñez, Raven Luther, Emily Huang, Emily Guzman, Carolina Arguelles-Grande, Kimon V. Argyropoulos, Margaret Black, Antonio Serrano, Melissa E. Call, Min Jae Kim, Brendan Belovarac, Tatyana Gindin, Andrew Lytle, Jared Pinnell, Theodore Vougiouklakis, John Chen, Lawrence H. Lin, Amy Rapkiewicz, Vanessa Raabe, Marie I. Samanovic, George Jour, Iman Osman, Maria Aguero-Rosenfeld, Mark J. Mulligan, Erik M. Volz, Paolo Cotzia, Matija Snuderl, Adriana Heguy

**Affiliations:** Institute for Systems Genetics, NYU Grossman School of Medicine, New York, USA; Department of Pathology, NYU Grossman School of Medicine, New York, USA; Genome Technology Center, Division of Advanced Research Technologies, Office of Science and Research, NYU Langone Health, New York, USA; Center for Biospecimen Research and Development, NYU Langone Health, New York, USA; MRC Centre for Global Infectious Disease Analysis and Department of Infectious Disease Epidemiology, Imperial College London; Department of Dermatology, NYU Grossman School of Medicine, New York, USA; Division of Infectious Diseases and Immunology, Department of Medicine and NYU Langone Vaccine Center, NYU Grossman School of Medicine, New York, USA; Department of Medicine, NYU Grossman School of Medicine, New York, USA; Medical Center IT, NYU Langone Health, New York, USA

## Abstract

Effective public response to a pandemic relies upon accurate measurement of the extent and dynamics of an outbreak. Viral genome sequencing has emerged as a powerful approach to link seemingly unrelated cases, and large-scale sequencing surveillance can inform on critical epidemiological parameters. Here, we report the analysis of 864 SARS-CoV-2 sequences from cases in the New York City metropolitan area during the COVID-19 outbreak in Spring 2020. The majority of cases had no recent travel history or known exposure, and genetically linked cases were spread throughout the region. Comparison to global viral sequences showed that early transmission was most linked to cases from Europe. Our data are consistent with numerous seeds from multiple sources and a prolonged period of unrecognized community spreading. This work highlights the complementary role of genomic surveillance in addition to traditional epidemiological indicators.

## Introduction

In December of 2019, the novel pneumonia COVID-19 emerged in the city of Wuhan, in Hubei province, China. Shotgun metagenomics rapidly identified the new pathogen as SARS-CoV-2, a betacoronavirus related to the etiological agent of the 2002 SARS outbreak (SARS-CoV), and of possible bat origin (Zhou et al. 2020; Andersen et al. 2020). Building on infrastructure from past outbreaks (Park et al. 2015; Carroll et al. 2015), genomic epidemiology has been applied to track the worldwide spread of SARS-CoV-2 using mutations in viral genomes to link otherwise unrelated infections (Grubaugh et al. 2019; Zhang and Holmes 2020). Collaborative development of targeted sequencing protocols (Quick et al. 2017; ARTIC Network), open sharing of sequences through the GISAID (Global Initiative on Sharing All Influenza Data) repository (Shu and McCauley 2017), and rapid analysis and visualization of viral phylogenies using Nextstrain (Hadfield et al. 2018) have provided unprecedented and timely insights into the spread of the pandemic. Notably, community transmission was identified using surveillance sequencing in the Seattle area in time to implement preventative measures (Bedford et al. 2020; Worobey et al. 2020).

The New York City metropolitan region rapidly became an epicenter of the pandemic following the identification of the first community-acquired case on March 3, 2020 (a resident of New Rochelle in nearby Westchester County who worked in Manhattan). As of May 10, 2020, New York State had 337,055 cases – the highest in the United States, and 8% of the worldwide total. Fully 55% of NY State cases lay within the five boroughs of New York City (185,357 cases), followed by Nassau and Suffolk counties to the east on Long Island (75,248 cases) (NYS Department of Health). The outlying boroughs and suburban counties reported markedly higher infection rates than Manhattan. The outbreak overlaps with the catchment area of the NYU Langone Health (NYULH) hospital system, including hospitals on the east side of Manhattan (Tisch/Kimmel), Brooklyn (formerly Lutheran Hospital), and Nassau County (Winthrop). Since even early COVID-19 cases presented mostly without travel history to countries with existing outbreaks, determining the extent of asymptomatic community spread and transmission paths became paramount. In parallel with increased clinical capacity for diagnostic PCR based testing, we sought to trace the origin of NYULH-treated COVID-19 cases using phylogenetic analysis to compare to previously deposited COVID-19 viral sequences. We further aimed to develop an approach to integrate sequencing as a complementary epidemiological indicator of outbreak trajectory.

## Results

To assess the spread of SARS-CoV-2 within the NYU Langone Health COVID-19 inpatient and outpatient population, we deployed and optimized a viral sequencing, quality control, and analysis pipeline by repurposing existing genomics infrastructure. Samples were selected for sequencing from those confirmed positive between March 12 and May 10, 2020. During this period, positive tests within the NYULH system mirrored those of New York City and nearby counties (**Supplemental Fig. 1**) (Petrilli et al. 2020). Illumina RNA-seq libraries were generated using a ribodepletion strategy starting from total RNA from nasopharyngeal swabs. Hybridization capture with custom biotinylated baits was used to enrich RNA-seq libraries for viral cDNA for sequencing (**Supplemental Fig. 2**, **Methods**). Of 1,113 libraries generated and sequenced, fully 78% yielded a sequence passing quality control (QC, see Methods). Pass rates were lower for samples with qRT-PCR Ct values > 30 (**Supplemental Fig. 3A-B**). We observed that high-quality sequences could be generated directly from shotgun libraries for qPCR Ct values < 30, thereby simplifying pooling and logistical constraints by skipping the capture step. Up to 23 samples were multiplexed in a single capture pool (**Supplemental Fig. 3C-D**). Samples with similar Ct values were grouped to minimize the range of target cDNA representation across a single capture pool (**Supplemental Fig. 3E-F**). Our pipeline was verified using a positive control synthetic RNA spiked in to total human RNA, as well as a negative water control (Methods). This resulted in 864 sequences passing quality control, representing 10% of COVID-19 positive cases in NYULH over that time period (**Supplemental Fig. 1, Supplemental Table 1**).

The cohort of 864 sequenced cases included a range of ages (**Fig. 1A**). Cases originated throughout the NYULH system, which comprises hospitals in the New York City boroughs of Manhattan and Brooklyn, and Nassau County, a suburb to the east of the city on Long Island (**Fig. 1B**). 66% of cases resided within New York City, and 86% within NY State (**Fig. 1C**). Analysis of residential ZIP codes showed that cases reflected the hospital catchment area within the New York metropolitan region (**Fig. 1D**). Our dataset included few cases from Westchester County to the north of the city where the earliest detected regional outbreak was concentrated, as it is outside of the NYULH catchment area.

We compiled a database for 820 of these cases from electronic medical records, including potential exposure information for health care worker status, travel history, and close contact with a COVID-19 individual (Methods). We found no recorded exposures for 43% of cases (**Fig. 1E**). Travel history was present in only 5% of cases, and these cases were concentrated in March (**Fig. 1F**). Of 14 cases where travel destination information was available, 9 destinations were within the US, 4 were in Europe, and 1 was in South Asia. This assessment relies upon clinical notes during a period where clinical capacity was stretched, thus likely underestimates potential exposures. Conversely, the potential exposure may have been coincidental given the uncontrolled community spread at the time.

**Figure 1.**
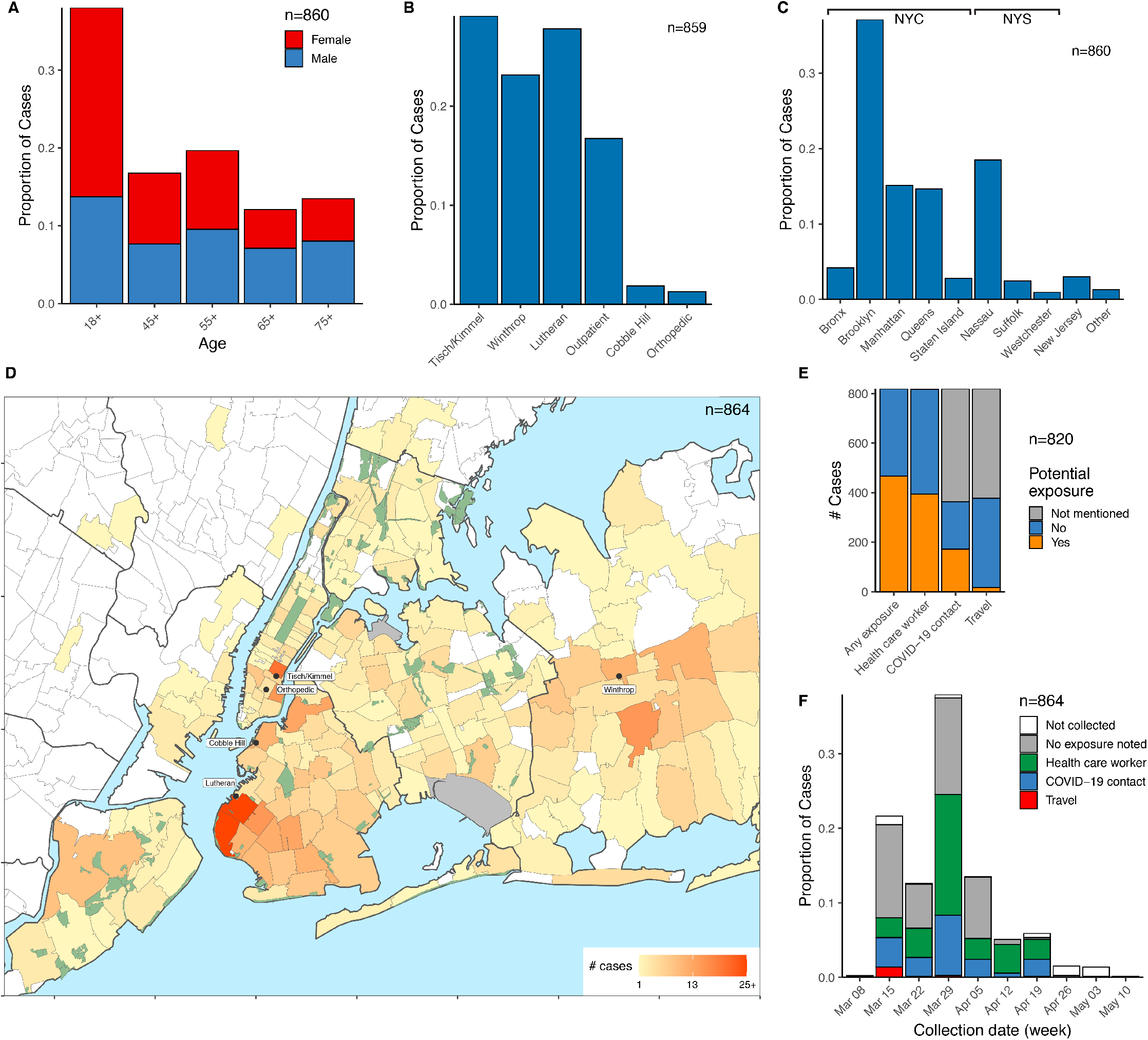
Demographic parameters of sequenced SARS-CoV-2 cases in the NYULH system. Cases are broken down by: (A) Age and sex. (B) Collecting hospital (C) Residential location, grouped by borough and outlying counties; “Other” includes counties with few cases. (D) Localization of case residences within the New York City Region. The color scale indicates numbers of cases per ZIP code. Collecting hospitals are indicated in rounded boxes. (E). Potential exposure status, categorized by occupation as healthcare worker, travel history, and contact with a COVID-19 positive individual. (F) Potential exposure status by collection date.

We inferred a maximum likelihood phylogeny to assess relatedness among cases (**Fig. 2**). Coloring cases by county of residence within the New York region showed identical or related viral sequences found across multiple counties from the onset of our sampling (**Fig. 2**). We detected 890 nucleotide and 547 amino acid mutations across all cases (**Supplemental Fig. 4**). Mutation of D614G in the spike protein, which has been suggested to affect transmission or virulence (Zhang et al. 2020), was present in >95% of sequences. Functional analysis will be required to determine whether functional changes can be ascribed to any of these mutations, and what role mutations might play in shaping the ongoing pandemic.

**Figure 2.**
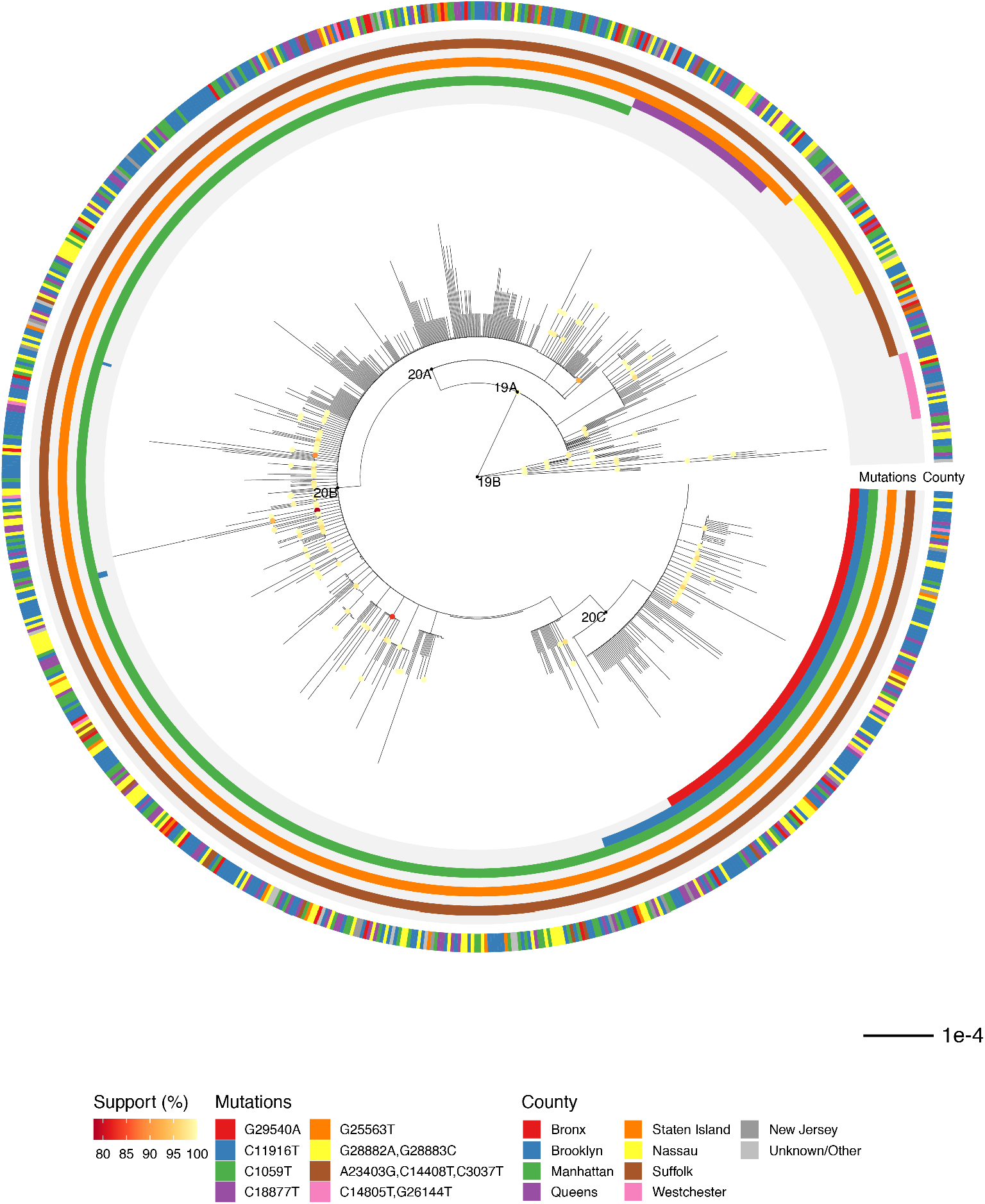
Phylogenetic relationship of regional viral sequences. Maximum likelihood phylogeny inferred from 864 cases. Nodes with bootstrap support values above 75 are colored. Inner rings indicate groups of clade-defining mutations. Outer ring indicates county of residence. Scale bar represents nucleotide substitutions per site.

We then assessed the relatedness of our cases to 5,004 sequences from across the world from the GISAID EpiCov repository (**Supplemental Fig. 5**). A maximum likelihood tree showed that cases from the NY region demonstrated broader diversity than initially reported in Seattle (Bedford et al. 2020), the only other US region with a comparable level of viral sequences (**Supplemental Fig. 6**). To investigate the timing of introductions to New York City, we inferred a rooted time-scaled phylogeny (**Fig. 3A, Supplemental Fig. 7A**). Analysis of our cases within this phylogeny identified 109 genotypes introduced to the northeast US (**Fig. 3B, Supplemental Table 3**). Identification of source nodes ancestral to at least one sequence from outside the northeast US in addition to these transmission chains placed most introductions broadly in late February and early March, slightly earlier than the first detected transmissions within New York City (**Fig. 3C, Supplemental Fig. 7B**). The timing of these introductions did not differ substantially under alternative nucleotide substitution models or rates (**Supplemental Fig. 7C**). The number of samples in each transmission chain varied widely, and two early transmission chains each comprised over three hundred cases. Only a minority of transmission chains included samples from Asia, while samples from Europe and the rest of the US were well-represented (**Fig. 3D**).

**Figure 3.**
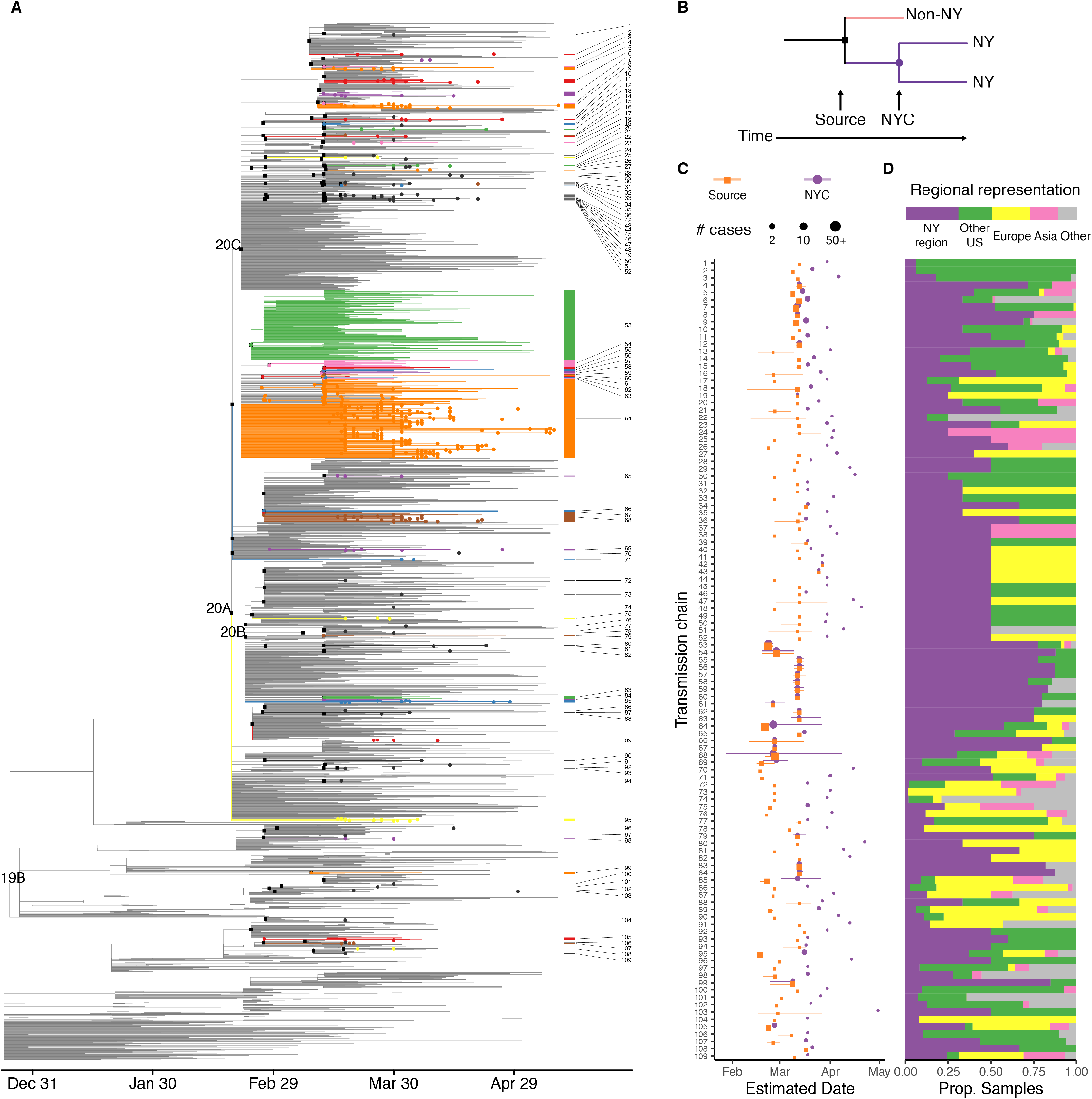
Time-scaled phylogeny showing global sequence context. (A) Colored edges highlight transmission chains. Black squares indicate source nodes, dots indicate detected presence in the northeast US. (B) Schematic of approach to infer introductions and transmission chains. (C-D) Transmission chains in the New York City region ordered by inferred divergence date from source. (C) Dates estimated for source transmission (orange) and earliest detected local transmission (purple) inferred from sequenced cases; lines represent 90% confidence intervals. Point size corresponds to the number of strains under source and all transmission chains. (D) Representation of global regions in each source transmission. Bar at top shows overall representation of regions in the phylogeny.

To assess the ongoing trajectory of the outbreak, we applied phylodynamic analysis to estimate viral effective population size from a subsample of sequences (Pybus and Rambaut 2009) (Methods). Under moderate assumptions, effective population size will be proportional to epidemic prevalence and growth rates of effective population size will correspond to epidemic growth (Volz et al. 2013). This analysis identified a period of rapid growth, followed by return nearly to the start point (**Fig. 4A**). We estimate that the peak effective population size occured on March 29 [95% CI: March 19-April 5]. The growth rate decreased steadily after March 1 and was negative with high confidence by mid-April (**Fig. 4B**), consistent with the epidemic curve of confirmed infections in the New York City Region (**Supplemental Fig. 1A**).

**Figure 4.**
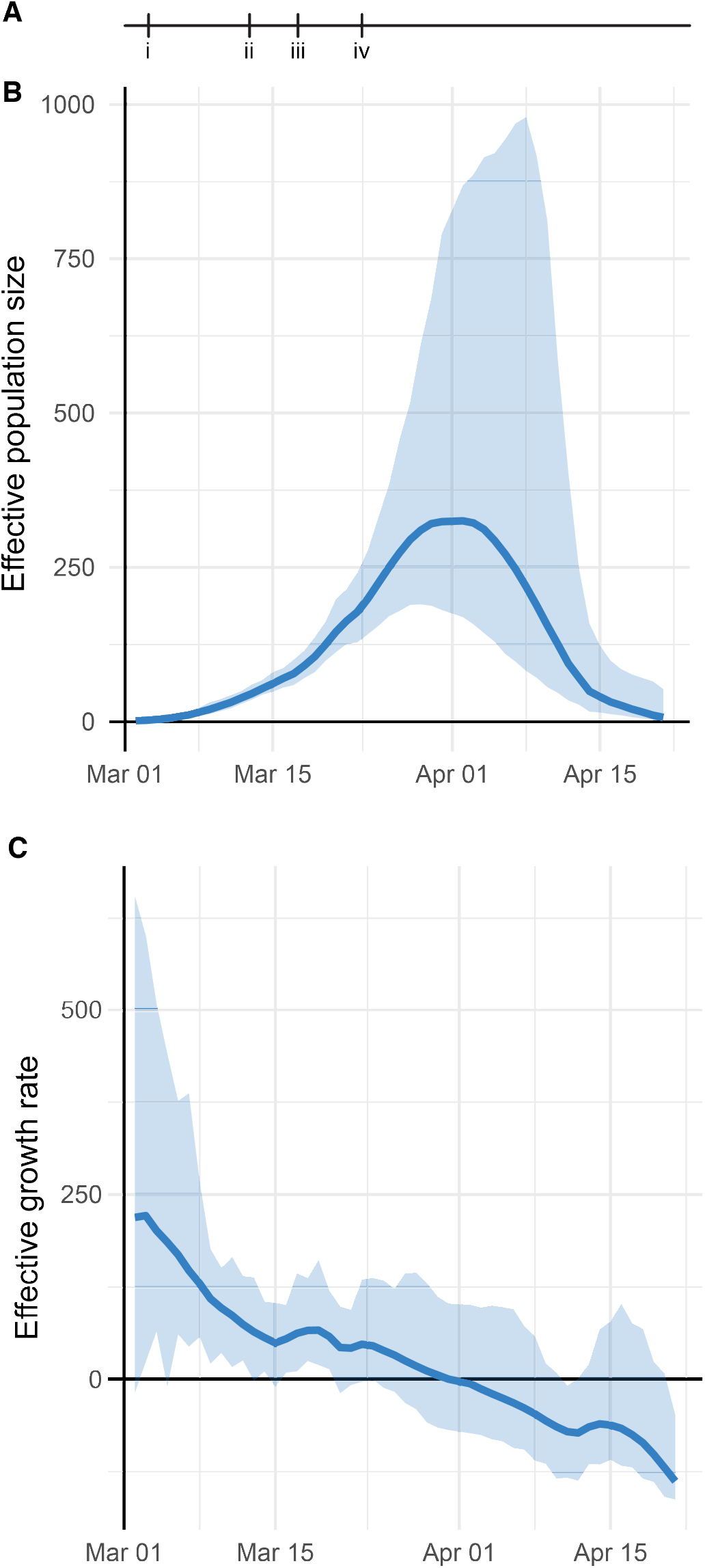
Phylodynamic analysis of outbreak trajectory. (A) Timeline of New York City outbreak, highlighting (i) announcement of first community-acquired case (Mar 3); (ii) ban on gatherings exceeding 500 people (Mar 12); (iii) closure of schools, restaurants, and bars, and other venues (Mar 16); (iv) closure of non-essential businesses (Mar 22). (B-C) Outbreak trajectory estimated from genetic data showing b. effective population size relative to March 1 and (C) growth rate of effective population size (units of 1/years). Shaded regions represent 95% credible interval.

## Discussion

Our work documents the genomic epidemiology of the COVID-19 outbreak in the New York City region in the Spring of 2020. The genetic data suggest that the New York outbreak was seeded by mid-February, and largely by way of Europe, which can be placed within the context of reduced travel flows from Asia to the US, the earlier spread of the pandemic from Asia to Europe, and the low overall prevalence in rest of the US. Several other reports of the initial stages of the New York City Region outbreak have identified early community spread on a similar timeframe (Gonzalez-Reiche et al. 2020; Fauver et al. 2020; Davis et al. 2020).

It is important to caution that fine-scale delineation of individual introductions and transmissions through genomic epidemiology is limited by viral mutation rate, incomplete sampling, and incomplete availability of exposure history (Villabona-Arenas et al. 2020). In particular, many early sequences demonstrate identical genotypes which could be consistent with additional transmission events, possibly by way of unsampled regions. Although our estimate of 109 introductions is thus likely to underestimate the total number of introductions, the genomic data are sufficiently informative to outline an unrecognized early spread in February which enabled rapid development of the outbreak in March. Further analysis (Worobey et al. 2020) and sequencing of archival samples will be needed to refine assessments of the initial spread.

Our demonstration of rapid sample processing, deposition, and analysis underscores the potential for genomic epidemiology to provide an independent estimate of disease transmission, and its potential to recognize impending resurgence of a regional outbreak. Further surveillance by medical centers, regional public health departments, and national efforts will be needed to monitor genomic epidemiology, pandemic spread, and public responses (**Supplemental Fig. 5**). Given the logistical, regulatory, and methodological challenges to establishing such surveillance during an outbreak, it is critical to have this infrastructure already in place (Kim et al. 2020) for future waves of COVID-19 or other future pandemics.

## Methods

### Bioethics statement

The collection of COVID-19 human biospecimens for research has been approved by NYU Langone Health (NYULH) Institutional Review Board under the S16-00122 Universal Mechanism of human bio-specimen collection and storage for research.

The approved IRB protocol allows for the collection and analysis of clinical, travel, exposure and demographic data (Osman et al. 2020). Electronic medical records were reviewed to compile a clinical database for 820 cases listing health care worker status, travel history, and close contact with a known COVID-19 case. For cases where a given exposure was not directly stated in the clinical record, we recorded that field as missing data but included other exposures in our analysis. A summary field of exposure history per case was generated by the presence of COVID-19 contact, travel history, or health care worker status, in that order.

### Sample collection

All samples were collected as part of clinical diagnostics. Nasopharyngeal swabs were collected and placed in 3 mL of Viral Transport Medium (VTM, Copan Universal Transport Medium) following clinical protocols. Samples were transported to the clinical microbiology laboratory at room temperature and tested for SARS-CoV-2 the same day. Remnant samples were stored at −70 °C.

### Clinical testing

All initial detection of COVID-19 cases was performed as part of the clinical care. Clinical testing was performed using the following three FDA Emergency Use Authorization (EUA) approved COVID-19 PCR based tests:

i. NYULH-validated PCR test using the US CDC primer design, targeting three regions of the virus nucleocapsid (N) gene, and an internal control primer targeting the human RNAse P gene (RP) (https://www.cdc.gov/coronavirus/2019-ncov/lab/rt-pcr-panel-primer-probes.html) with PCR carried out on an ABI7500 Dx system. The limit of detection is 10,000 copies/mL.
ii. The Roche Cobas 6800 RT-PCR platform targeting the Orf1/a and E sequences, per manufacturer instructions. The limit of detection is 180 copies/mL.
iii. The Cepheid Xpert Xpress RT-PCR platform targeting the N2 and E viral sequences, per manufacturer instructions. The limit of detection is 250 copies/mL.

### RNA extraction

RNA extraction was performed using two platforms for parallel sample processing:

i. Using the Maxwell RSC instrument (Promega, cat. AS4500), total RNA was extracted from 300 *μ*L of viral transport medium with the buccal swab DNA kit (Promega, cat. AS1640). The following modifications were introduced to extract total RNA as opposed to total nucleic acids: samples were incubated at 65 °C for 30 min for proteinase K digestion and virus deactivation, and DNase I (Promega) was added to the reagents cartridge to remove genomic DNA during nucleic acids extraction. Total RNA was eluted in 50 *μ*L of nuclease-free water.
ii. Using the KingFisher Flex System (ThermoFisher Scientific) system, RNA was extracted from heat-inactivated nasopharyngeal swab samples in batches of 96 samples, following the manufacturer’s instructions and the MagMax *mirVana* Total RNA isolation Kit (ThermoFisher Scientific, A27828). Briefly, 250 uL of nasopharyngeal swab collection was lysed in lysis buffer and β-mercaptoethanol and subsequently bound to magnetic beads and loaded into the KingFisher Flex instrument. A DNase I treatment step was performed as part of the instrument protocol and RNA samples were eluted in 50 uL of Elution Buffer and immediately stored at −80C.

### Library preparation and sequencing

Illumina sequencing libraries were prepared from 10 *μ*L of total RNA. Two methods for cDNA RNA-seq library preps were used, both based on a ribodepletion approach:

i. KAPA RNA HyperPrep Kit with RiboErase (HMR) (Roche Kapa cat. KK8561). We followed the manufacturer’s protocol, with the following modifications: for the adapter ligation step, we prepared a plate of barcoded adapters (IDT) at a concentration of 500 nM, and performed 15 cycles of PCR amplification of the final library.
ii. Nugen Trio with human rRNA depletion (Tecan Genomics, cat. 0606-96), including DNase I treatment, cDNA synthesis, single primer isothermal amplification (SPIA), enzymatic fragmentation, library construction, final PCR amplification (12-16 cycles), and an AnyDeplete step to remove host rRNA transcripts. An automated protocol was implemented on a Biomek FX^P^ liquid handler integrated with a Biometra TRobot 96-well thermal cycler (Beckman Coulter).

Purified libraries were quantified using qPCR (Kapa Biosystems, KK4824). Library size distribution was checked using an Agilent TapeStation 2200.

Libraries presumed more suitable for capture (generally, qPCR Ct value > 30) were enriched for SARS-CoV-2 genomic sequences using custom biotinylated DNA probe pools either from Twist Biosciences or Integrated DNA Technologies:

i. For capture using the IDT xGen COVID capture panel (Integrated DNA Technologies, cat. 10006764), we followed the manufacturer’s protocol. Briefly, hybridization of 500 ng to 1 *μ*g of combined library DNA with 4 *μ*L of XGen Lockdown probes was carried out at 65 °C for 4-16 h, followed by PCR amplification for 6-10 cycles.
ii. For capture using the Twist Bioscience custom panel (Twist Design ID: TE-95888003, generously shared by the Seattle Flu Study), we followed the manufacturer’s protocol using the Twist Hybridization and Wash Kit (Twist Biosciences, cat. 101025). Hybridization of 1-2 *μ*g combined library DNA was carried out at 70 °C for 16-20 h. Post-capture PCR amplification cycles ranged from 12-14 cycles.

In general, we pooled samples with similar Ct values and accounted for variations in parent library concentration, multiplexing up to 23 libraries per reaction.

Samples were sequenced as paired end 100- or 150-cycle reads on the NextSeq 500 or NovaSeq 6000 (using SP or S1 flow cells). All flow cells were loaded such that indexing barcode sequences for multiplexed samples differed by 3 bp or more.

Control samples were processed as follows: a positive control of 5 ng of synthetic viral RNA (Genbank MT007544.1, Twist Bioscience) was spiked into human total RNA (ThermoFisher cat. 4307281). A negative control sample was generated from H2O put into RNA extraction.

### Sequenced read processing

Reads were demultiplexed with Illumina bcl2fastq v2.20 requiring a perfect match to indexing barcode sequences. All RNA-seq and Capture-seq data were processed using a uniform mapping pipeline. Illumina sequencing adapters were trimmed with Trimmomatic v0.39 (Bolger et al. 2014). Reads were aligned using BWA v0.7.17 (Li and Durbin 2009) to a custom index containing human genome reference (GRCh38/hg38) including unscaffolded contigs and alternate references plus the reference SARS-CoV-2 genome (NC_045512.2, wuhCor1). Presumed PCR duplicates were marked using samblaster v0.1.24 (Faust and Hall 2014). Only sequences with >23000 bp unmasked sequence were analyzed. Duplicate sequences from the same case were excluded, resulting in 864 final sequences (**Supplemental Table 1**). Variants were called across all samples using bcftools v1.9:

> bcftools mpileup —redo-BAQ --adjust-MQ 50 —gap-frac 0.05 —max- depth 100 00 --max-idepth 2 0 00 00 --output-type u |
>
> bcftools call --ploidy 1 —keep-alts —multiallelic-caller -f GQ

Raw pileups were filtered using

> bcftools norm --check-ref w --output-type u |
>
> bcftools filter -i “INFO/DP>=10 & QUAL>=10 & GQ>=99 & FORMAT/DP>=10” --SnpGap 3 —IndelGap 10 —set-GTs. —output-type u |
>
> bcftools view -i ‘GT=“alt”’ --trim-alt-alleles

Viral sequences were generated by applying VCF files to the reference sequence using ‘**bcftools consensus**’ with -m to mask sites below 20x with Ns, and -m N to mask sites of ambiguous genotypes with N.

### Geoplotting

The regional case heat map was generated using R v3.6.2 using the packages ggplot2 v3.3.0 for plotting, and sf v0.8 for geospatial data manipulation. Maps were generated based on the 2018 ZIP code tabulated area geographical boundaries obtained from the United States Census Bureau (United States Census Bureau).

### Phylogenetic analysis

Sequences for non-NYULH cases were downloaded from GISAID EpiCov on June 14, 2020 and filtered to sequences collected on or before May 10, 2020. Sequences from non-human hosts, annotated by Nextstrain as duplicate individuals or highly divergent, with <27,000 non-ambiguous nucleotides, or with improperly formatted dates or location were excluded. Sequences from outside New York state were subsampled to a maximum of 20 samples per admin division (US) or country (outside US) per month, prioritizing sequences most similar to the focal set of 864 NYULH samples. This priority was penalized if many non-US samples were most similar to the same US sample, and mutations were weighted 333x more heavily than masked sites. Global sequences were then combined with the sequences from this study.

Sequences were analyzed using the augur v7.0.2 pipeline (Hadfield et al. 2018). Sequences were aligned along with the reference genome using MAFFT v7.453 (Katoh and Standley 2013), and the resulting alignment was masked to remove 100 bp from the beginning, 50 from the end, and uninformative point mutations (positions 11083, 13402, 21575, 24389, 24390).

Maximum likelihood phylogenetic reconstruction was performed with IQ-TREE v1.6.12 (Nguyen et al. 2015) using a GTR substitution model and the -czb option. Support values were generated with the ultrafast bootstrapping option with 1000 replicates. This tree was used to tabulate nucleotide and amino acid changes specific to lineages and cases; gaps with respect to the reference were reported as deletions. TreeTime v0.7.4 (Sagulenko et al. 2018) was used to generate a timetree rooted at the reference sequence, using the --keep-polytomies option, and under a strict mutational clock under a skyline coalescent prior with a rate of 8 × 10^-4^ mutations per site per year and a standard deviation of 4 × 10^-4^.

For each NYULH case, the inferred earliest New York City transmission was identified as the most ancestral node or tip with >70% of sequences originating in the northeast (NY, CT, NJ, PA) on the time-scaled phylogeny using the ape (Paradis and Schliep 2019) and phangorn (Schliep 2011) R packages. The transmission source was identified as the first ancestral node defined by a unique mutation and ancestral to a sequence originating outside the northeast. Transmissions with identical source nodes were grouped to yield transmission chains. Trees were plotted with the tidygraph and ggraph R packages.

### Phylodynamic Analysis

To minimize ascertainment and sampling bias, analysis was performed on a subset of sequenced cases residing in New York City and outlying Westchester, Nassau and Suffolk counties, and excluded outpatients and known health care workers. Sequence data were aligned to reference (accession NC_045512.2) and ends trimmed using MAFFT 7.450 (Katoh and Standley 2013). A maximum likelihood tree was estimated using IQ-TREE 1.6.1 using a HKY substitution model (Nguyen et al. 2015). A further 20 phylogenies were derived by randomly resolving polytomies and enforcing a small minimum branch length of 7×10^-6^ substitutions per site using the ape R package (Paradis and Schliep 2019). Rooted time scaled phylogenies were estimated using the treedater R package version 0.5.1 (Volz and Frost 2017) using a strict molecular clock. The skygrowth R package version 0.3.1 (Volz and Didelot 2018) was used to estimate effective population size through time with an exponential prior for the smoothing parameter with rate 10^-4^. The final estimates were generated by averaging results over the 20 estimated time trees. A script for reproducing these results is available at: https://gist.github.com/emvolz/d58cce01c3310a01df09faf615b77070.

## Data Availability

Raw sequencing reads filtered to remove the host genome are available from NCBI SRA under BioProject PRJNA650245. Sequences have been deposited into the GISAID repository immediately upon QC with virus name "NYUMC" and can be visualized at http://nextstrain.org/ncov.

http://www.mauranolab.org/SARSCoV2.html

## Software availability

Code used in data processing are available at https://github.com/mauranolab/mapping/tree/master/dnase.

## Acknowledgements

We are indebted to the NYULH clinicians and laboratory personnel involved in the care and testing of the patients in this study. We would like to thank all the laboratories who have contributed sequences to GISAID (**Supplemental Table 2**). We thank Lea Starita and the Seattle Flu Study for technical assistance and sharing their bait design. This work was partially funded by NIH grants R35GM119703 (M.T.M.), P50CA016087 (I.O. and G.J.), P30CA016087 (I.O., P.C., and A.H.), UM1AI148574 (M.J.M), the NYULH Office for Science and Research, and MR/R015600/1 from the MRC Centre for Global Infectious Disease Analysis, School of Public Health, Imperial College London (M-R.C., E.M.V.)

## Author contributions

M.T.M., M.J.M., P.C., M.S., and A.H. conceived and supervised the study. L.B., G.S., X.F., C.A-G., K.V.A., M.B., A.S., M.E.C., M.J.K., B.B., T.G., A.L., J.P., T.V., L.H.L., A.R., V.R., M.I.S., G.J., I.O., M.A-R., M.J.M, P.C., and M.S. collected clinical samples and data. S.R., P.Z., D.D., G.W., M.S.H., P.M., Y.Z., and A.H. generated sequencing data. C.M., J.Ca., E.G. and J.Ch. contributed informatics tools. M.T.M, A.M.R., N.A.V., M.S.H, M.R-C., L.G., R.O., R.L., E.H., E.M.V., and A.H. performed the data analysis. M.T.M, E.M.V., M.S., and A.H. wrote the manuscript.

---

**Supplemental Fig. 1.**
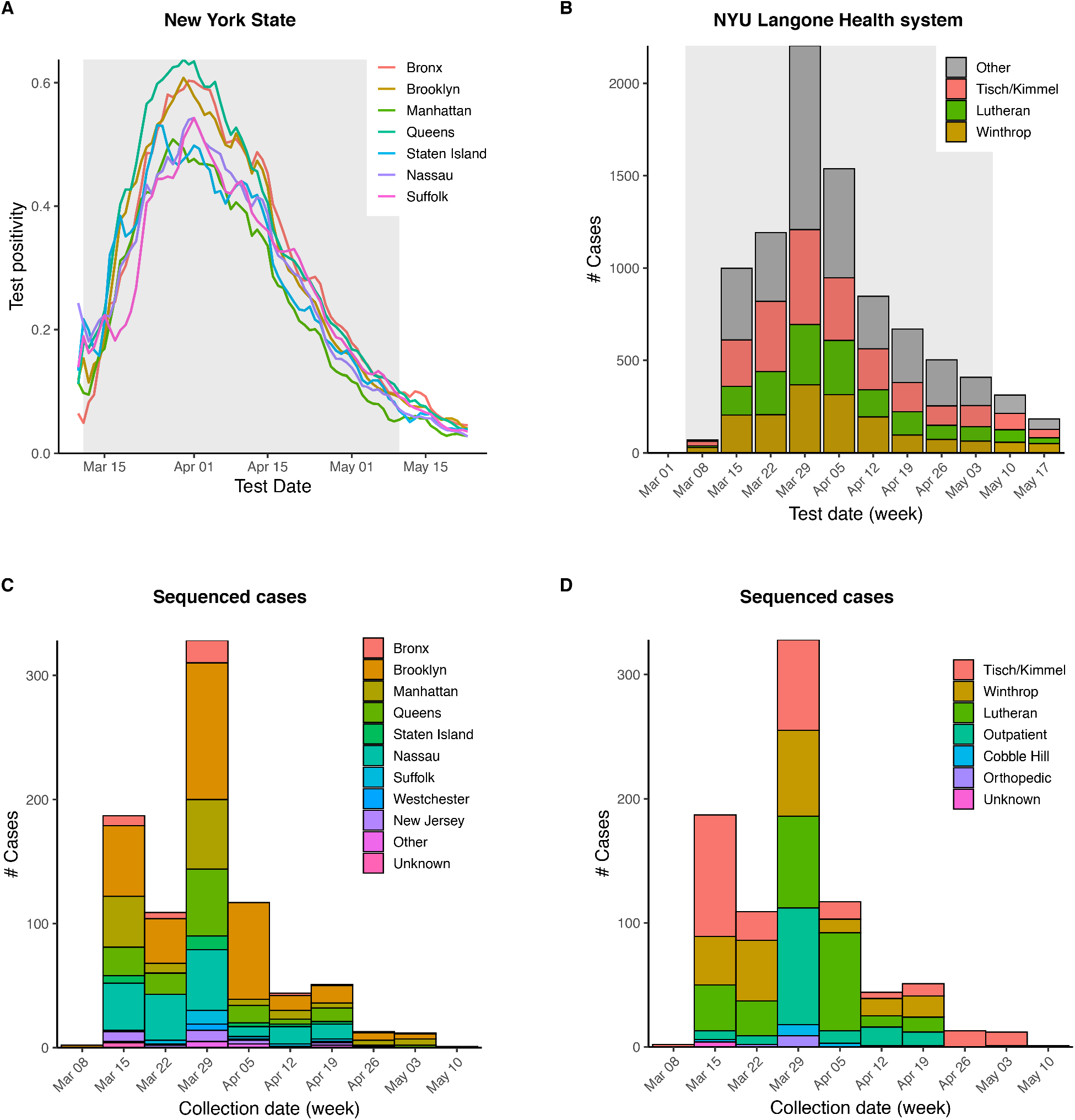
Outbreak trajectory and sampling of NYU Langone Health catchment area. (A) SARS-CoV-2 positivity rate for New York City boroughs and outlying counties reported by New York State Department of Health (NYS Department of Health). (B) Summary of weekly positive tests across NYU Langone Health. Shaded region indicates time period sampled for sequenced cases. (C-D) Sequenced cases by collection date, broken down by county of residence (C) and collecting hospital (D).

**Supplemental Fig. 2.**
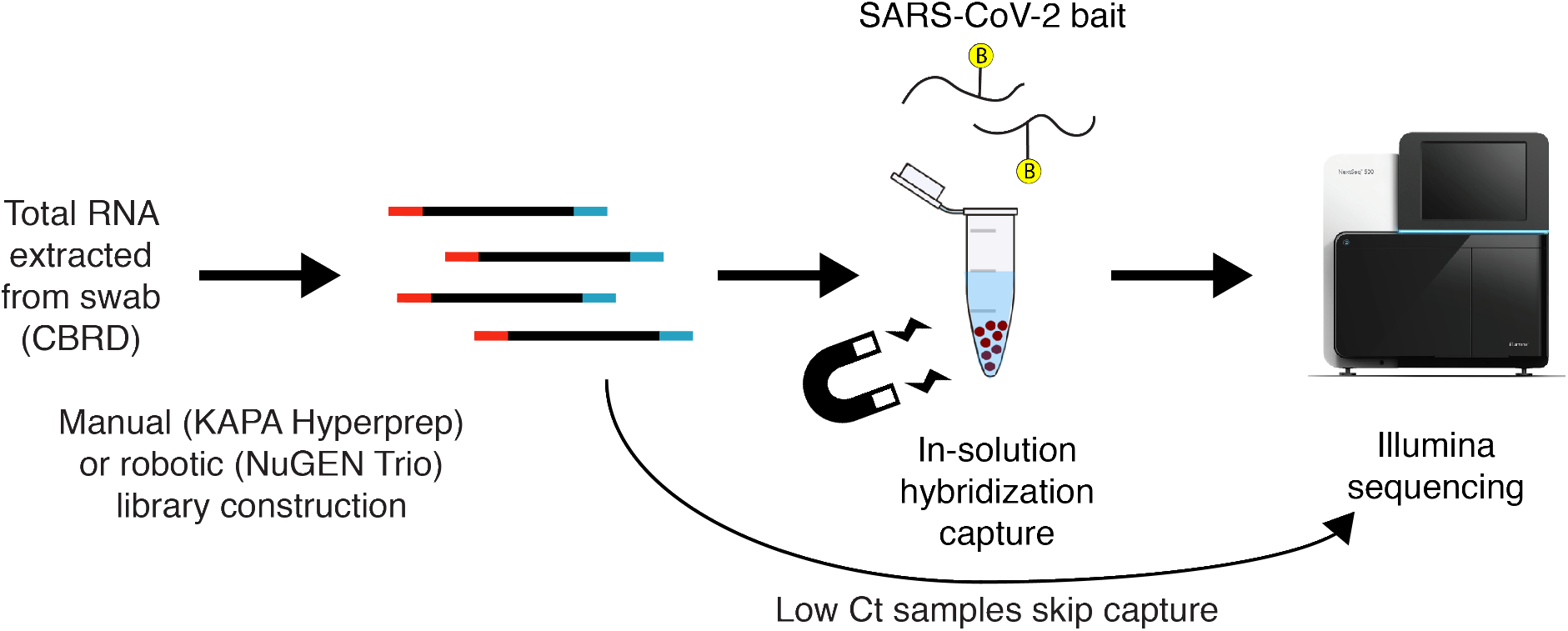
SARS-CoV-2 sequencing pipeline. Total RNA was extracted using high throughput extractors, at the Center for Biorepository Specimen and Development (CBRD) at NYU Langone Health. RNA-seq libraries were prepared using two ribodepletion protocols. For samples with qPCR Ct values < 30, we skipped the hybridization capture enrichment and sequenced RNA-seq libraries directly; other libraries were enriched for the viral genome sequences using hybridization-based capture baits (IDT or Twist) designed against the Wuhan SARS-CoV-2 RefSeq. Libraries were sequenced on the Illumina NovaSeq 6000 or NextSeq 500.

**Supplemental Fig. 3.**
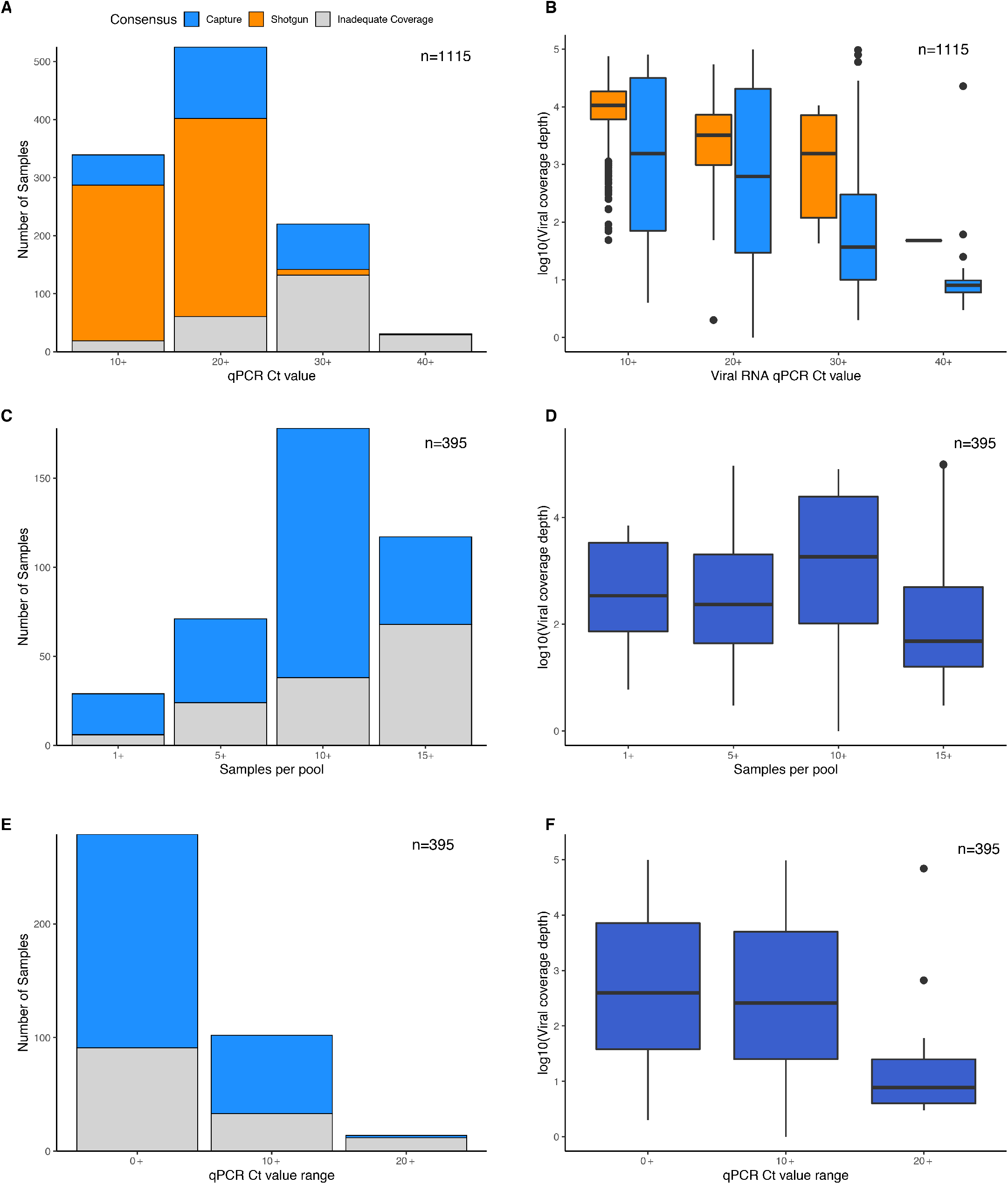
Technical factors related to sequencing quality control. Shown are sample counts by final quality control (QC) outcome (left) and average coverage depth (right) stratified by qRT-PCR Ct values (A-B), size of capture pool (C-D), and qRT-PCR Ct range among samples in the same capture pool (E-F). Boxes in B, D, F indicate first and third quartiles, whiskers extend to 1.5 times interquartile range.

**Supplemental Fig. 4.**
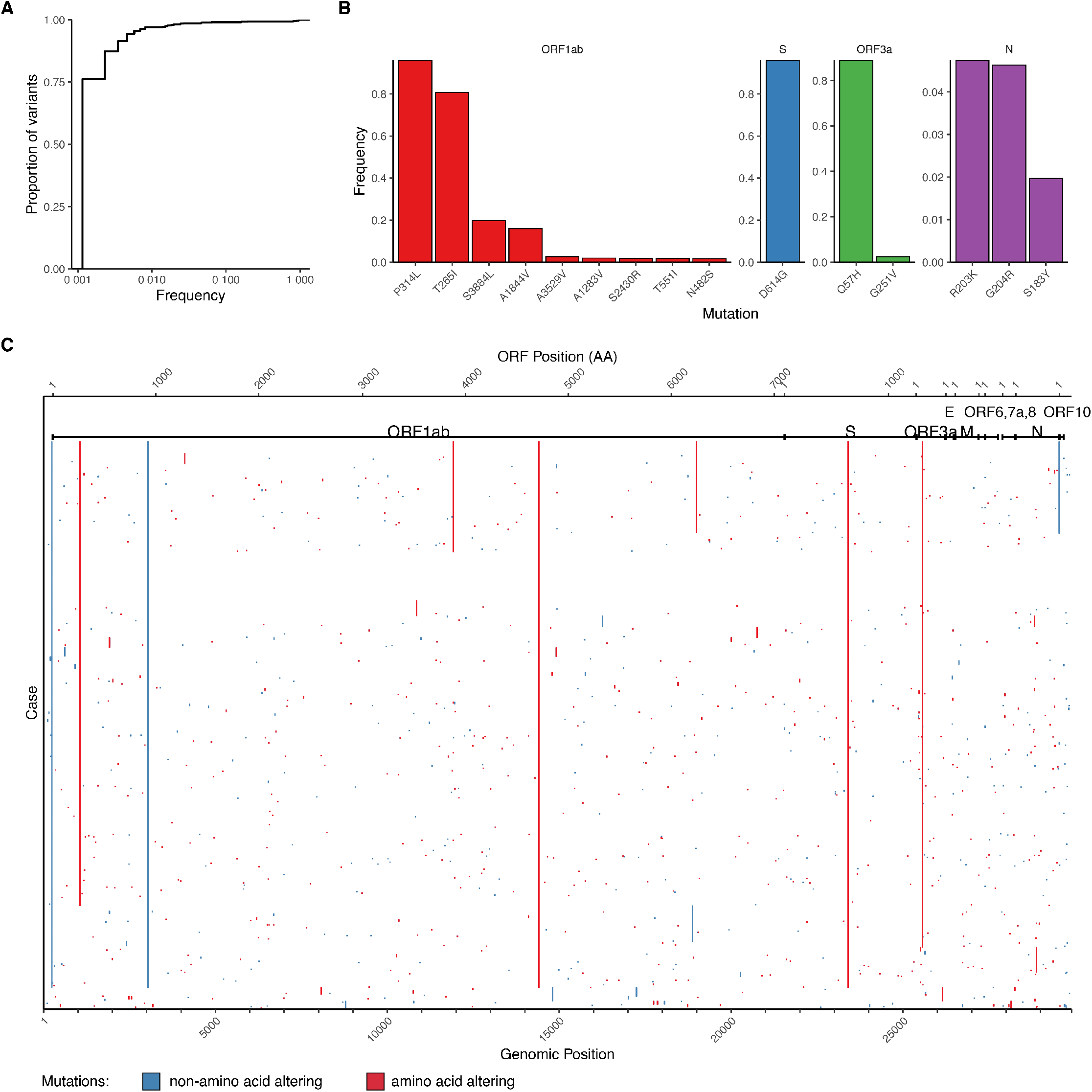
Overview of mutations identified. (A) Cumulative frequency distribution of variants identified across 864 cases. (B) Frequency for top 15 amino acid-altering mutations by open reading frame (ORF). (C) Heatmap of mutations identified per case, with x-axis being genomic coordinates, and rows in the same order as **Figure 2A**. ORFs are annotated according to GenBank MN908947.3.

**Supplemental Fig. 5.**
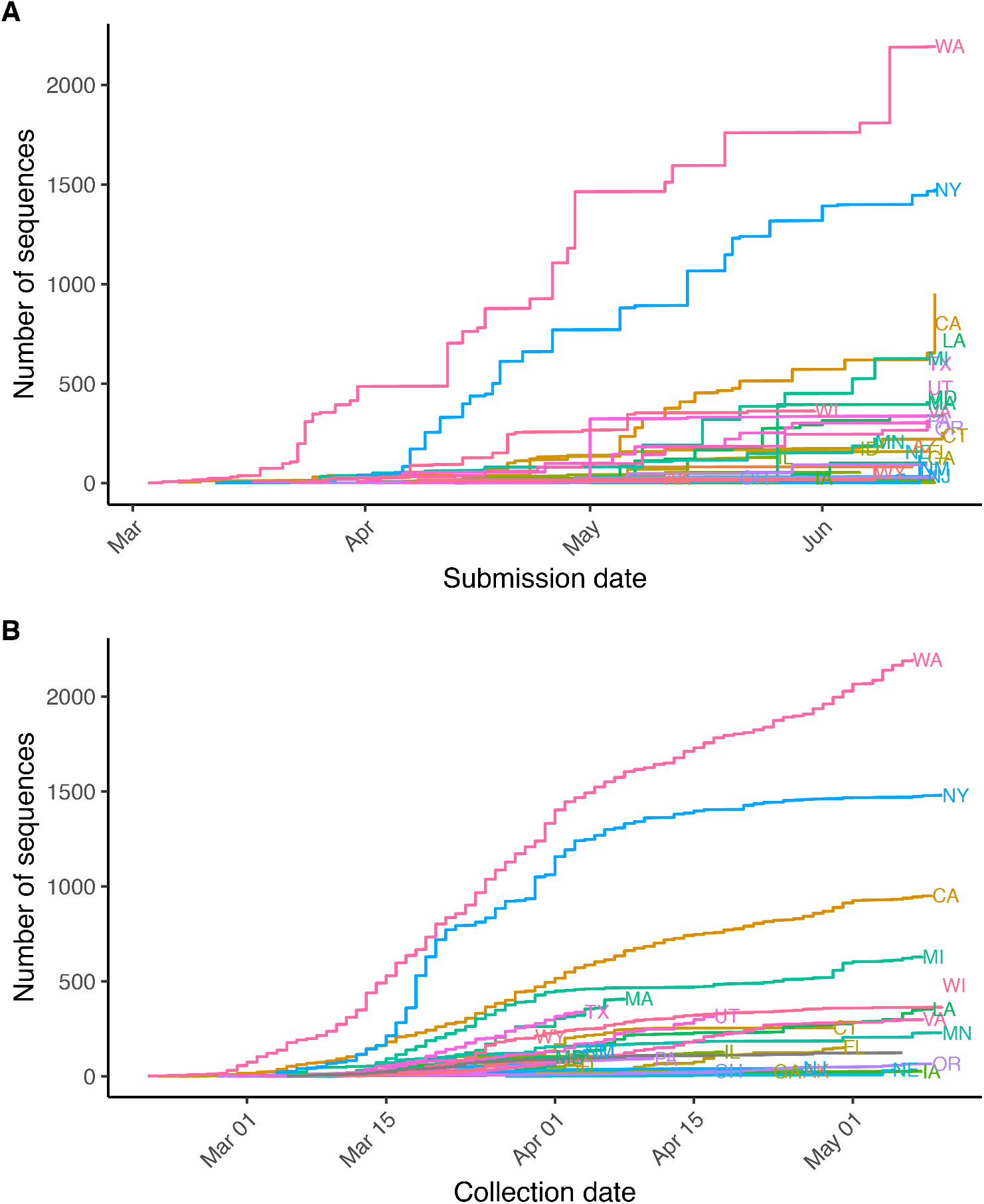
SARS-CoV-2 sequences in GISAID by US state. Summary of US sequences in the GISAID EpiCov repository collected through May 10, 2020 showing all submitters per state by (A) submission date, and (B) collection date.

**Supplemental Fig. 6.**
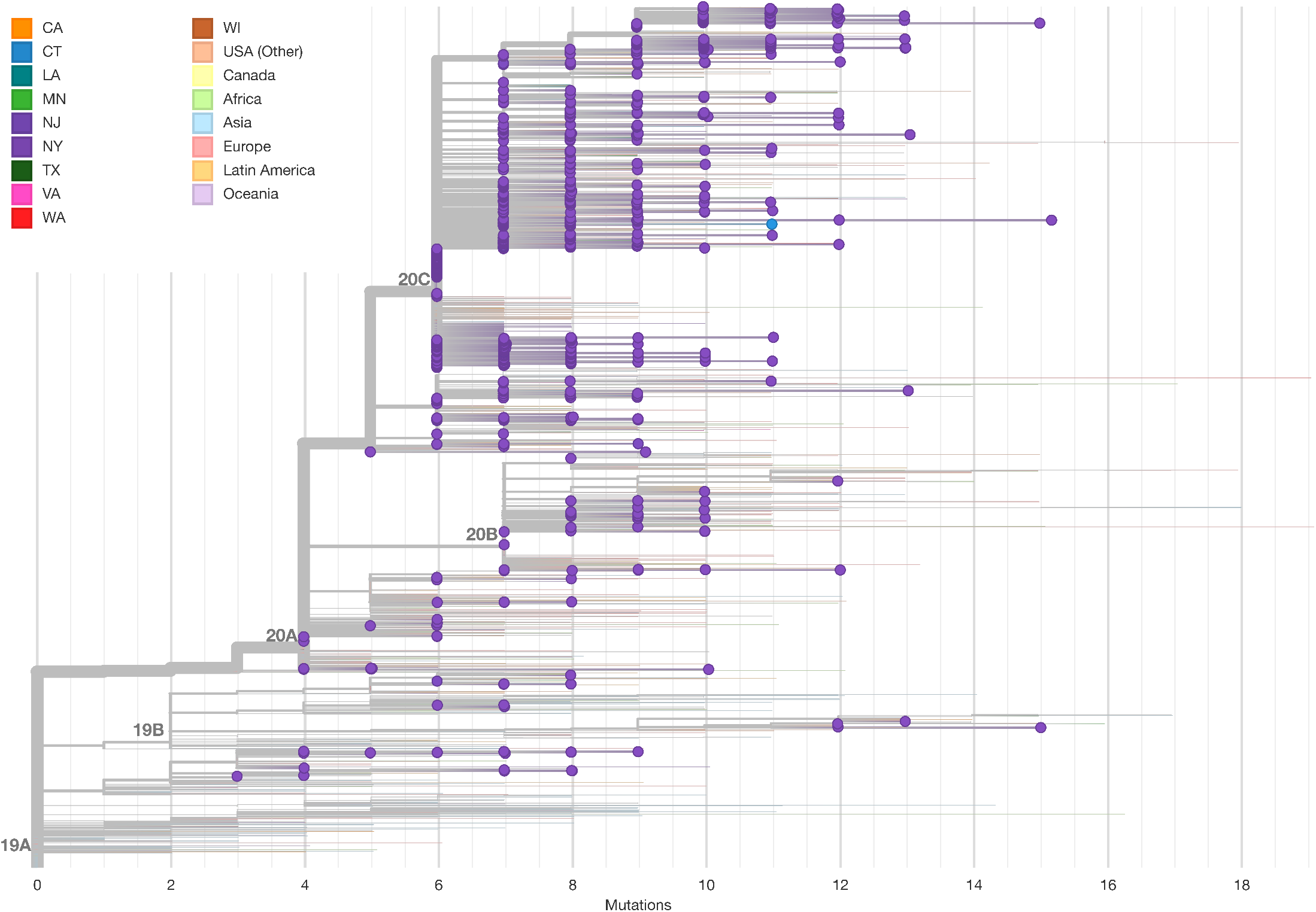
Maximum likelihood tree including 5,004 global sequences from GISAID. NYULH sequences are highlighted with dots, tip and edge coloring indicates geographical location.

**Supplemental Fig. 7.**
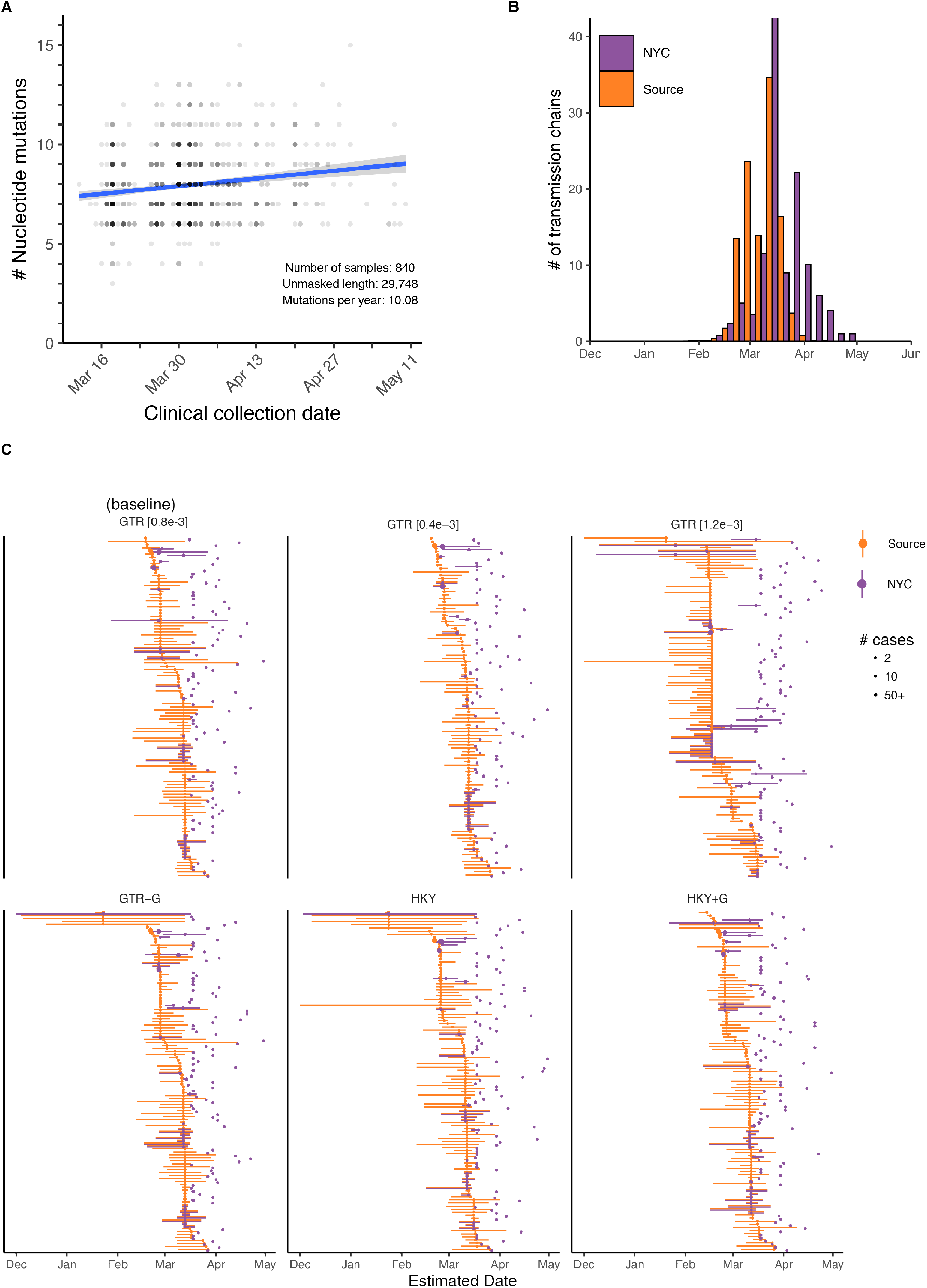
Time-scaled phylogeny analysis. (A) Root to tip plot showing the number of point mutations per case by collection date. Sequences with >10% ambiguous nucleotides are excluded. (B) Histogram of estimated dates for transmission chains identified in **Fig. 3**. (C) Effect of alternate substitution models (GTR+G, HKY and HKY+G) and substitution rates (0.4e-3 or 1.2e-3) on transmission chain dating. A time scaled phylogeny was inferred under alternate substitution models and substitution rates, and transmission chains re-identified for each.

## Supplemental Tables

Supplemental Tables are available as separate files.

**Supplemental Table 1. Summary of sequencing data**.

Per sample summary of sequencing data for 864 cases. PropViralReads, proportion of nonredundant reads mapping to SARS-CoV-2 genome; analyzedViralReads, number of reads mapping to SARS-CoV-2 genome and passing all filters; PropDupViralReads, proportion of analyzedViralReads marked as PCR duplicates; Mean_Viral_Coverage, mean coverage depth; Num_bp_20x, number of bp covered at ≥20x depth

**Supplemental Table 2. Acknowledgements of GISAID sequences used**.

List of sequences and contributors from GISAID.

**Supplemental Table 3. New York City Region transmission chains**.

Summary of 109 transmission chains, including strain genotypes and date of nodes representing divergence from source and first NYC transmission.

